# WNK2 facilitates ovarian cancer progression by upregulating POU5F1B

**DOI:** 10.1101/2025.08.26.25334517

**Authors:** Fengjie Li, Yongqin Jia, Xiaoli Min, Pangyang Zhang, Yudi Li, Deng li, Lanqin Cao, Yanzhou Wang, Zhiqing Liang

## Abstract

Ovarian cancer (OC) remains the most lethal gynecological malignancy. Our previous work established that WNK lysine-deficient protein kinase 2 (WNK2) promotes OC cell proliferation and migration. To elucidate how WNK2 drives OC progression, we performed transcriptome sequencing to identify WNK2-regulated mRNAs and noncoding RNAs. Candidate targets were validated via qRT-PCR and Western blot. Functional assays (CCK-8, colony formation, Transwell) assessed the role of POU5F1B and its ability to rescue WNK2 knockdown effects. Given AKT’s involvement downstream of POU5F1B, we measured AKT phosphorylation. Additionally, since WNK2 activates RAS (as previously shown), we tested whether RAS inhibition blocks WNK2-mediated POU5F1B regulation. POU5F1B exhibited oncogenic properties in OC cells. WNK2 upregulated POU5F1B mRNA and protein levels, and POU5F1B overexpression reversed tumor-suppressive effects caused by WNK2 knockdown. Mechanistically, WNK2 depletion reduced AKT phosphorylation, which was restored by POU5F1B overexpression. Furthermore, RAS inhibition abolished WNK2-driven POU5F1B upregulation, linking WNK2-RAS signaling to POU5F1B activation. Our study demonstrates that WNK2 promotes OC progression by upregulating POU5F1B, thereby activating AKT signaling. These findings solidify WNK2’s oncogenic role and highlight its therapeutic potential in OC.

## 1. Introduction

Ovarian cancer (OC) remains the most lethal malignancy of the female reproductive system. In the United States alone, 21,750 new OC cases were diagnosed in 2020, resulting in 13,940 deaths ^[1]^. High-grade serous ovarian carcinoma (HGSOC), the most aggressive histological subtype, accounts for 70-80% of OC cases and is responsible for the majority of OC-related mortality^[2]^.While primary treatment still relies on cytoreductive surgery and platinum-based chemotherapy, recent years have seen the emergence of novel therapeutic approaches. These include antiangiogenic agents (e.g., VEGF inhibitors), tyrosine kinase inhibitors, and folate receptor-targeted therapies ^[3]^. Notably, PARP inhibitors have revolutionized treatment for patients with BRCA1/2 mutations ^[4]^. However, therapeutic efficacy remains limited by drug resistance, toxicity, and the heterogeneous nature of OC ^[5]^. Consequently, the 5-year survival rate for advanced-stage OC has shown minimal improvement over the past decade, underscoring the urgent need to identify novel molecular targets for more effective therapeutic strategies.

WNK kinases, a family of serine/threonine kinases discovered in 2000, are distinguished by the absence of a conserved lysine in subdomain II . Initially recognized for their roles in blood pressure regulation and embryonic development ^[7, 8]^. WNK kinases have recently garnered significant attention for their functions in cancer. Among them, WNK2 has been identified as a tumor suppressor in multiple cancer types ^[9]^. For instance, its inactivation promotes glioblastoma growth and metastasis via Rac1 activation ^[10]^. WNK2 expression is reduced in meningiomas due to aberrant CpG island methylation and suppresses tumorigenesis by inhibiting colony formation ^[11]^. Similarly, WNK2 acts as a tumor suppressor in gastric, cervical, and pancreatic cancers by inactivating ERK signaling^[12–14]^. Intriguingly, our findings reveal a contrasting oncogenic role for WNK2 in ovarian cancer (OC) ^[15]^. The underlying reason for this dichotomous function of WNK2 in OC compared to its tumor-suppressive roles in cancers like glioma, gastric, colon, liver, and pancreatic cancer remains unclear. To investigate how WNK2 contributes to OC pathogenesis, we performed RNA-seq analysis to identify its transcriptional targets. This approach revealed that WNK2 specifically upregulates the expression of POU5F1B at both the mRNA and protein levels. POU5F1B, located on the cancer susceptibility locus chromosome 8q24 ^[16]^, encodes a membrane-enriched protein with 95% similarity to OCT4 ^[17]^. This transposon-activated gene promotes malignancy in colorectal cancer^[18]^ and is highly expressed in hepatocellular, breast, gastric, cervical, esophageal, and prostate cancers, where it correlates with poor survival and drives tumor progression ^[19–21]^. Adjacent to *Myc*—the most frequently mutated oncogene in OC—POU5F1B cooperates with it to promote cancer ^[22]^ and exerts its oncogenic effects partly by enhancing AKT activity ^[23]^. In this study, we establish POU5F1B as an oncogene in ovarian cancer (OC) and demonstrate that WNK2 promotes OC development by upregulating POU5F1B, thereby activating AKT. These findings provide mechanistic evidence for the tumor-promoting role of WNK2 in OC.

## 2. Materials and methods

### 2.1 Cell culture

Human ovarian cancer (OC) cell lines CAOV3 and A2780 were obtained from the American Type Culture Collection (ATCC, Manassas, VA, USA). Cells were cultured in Dulbecco’s Modified Eagle’s Medium (DMEM) supplemented with 10% fetal bovine serum (FBS) and 1% penicillin-streptomycin antibiotic solution (Gibco, Gaithersburg, MD, USA). Cultures were maintained at 37°C in a humidified atmosphere containing 5% CO₂.

### 2.2 Cell transfection and lentivirus transduction

The empty vector and POU5F1B-overexpression (POU5F1B-OE) plasmids were synthesized by Tsingke Biotechnology Co., Ltd. (Beijing, China). The WNK2-overexpression (WNK2-OE) plasmid and its corresponding empty vector were constructed by Genechem Co., Ltd. (Shanghai, China). siRNAs targeting WNK2 and POU5F1B were obtained from RiboBio (Guangzhou, China), with the following sequences: si-WNK2-1: CAA GGA CAA TGG AGC CATA; si-WNK2-2: GGA GTA TGC TAG GCT ATGA; and si-WNK2-3: CGA TGA AAT TGC CAC GTAT. si-POU5F1B-1: AGA AGT CCC AGG ACA TCAA, si-POU5F1B-2: CAC TGC AGA TCA GCC ACAT, and si-POU5F1B-3: CCC AGT CTC CGT CAT CACT. Transient Transfection: Cells were transfected 24 hours after plating. For each transfection, approximately 3 μg plasmid or 50 nM siRNA was mixed with 5 μL Lipofectamine 3000 reagent in serum-free medium. After 6 hours, the transfection mixture was replaced with complete medium containing serum.

Stable Cell Line Generation: To generate stable WNK2-knockdown cells, CAOV3 cells were transduced with sh-WNK2 lentiviral particles. Subsequently, these sh-WNK2 CAOV3 cells were transduced with POU5F1B-overexpression (POU5F1B-OE) lentivirus. Stable cell lines (sh-WNK2 + Vector and sh-WNK2 + POU5F1B-OE) were selected using 2 μg/mL puromycin ^[24]^.

### 2.3 Transcriptome sequencing analysis

Following WNK2 knockdown via siRNA transfection, total RNA was isolated using TRIzol™ reagent (Takara Bio, Inc.) with three biological replicates per condition. Differential expression analysis was performed by Beijing Novogene Institute (Beijing, China) using the DESeq2 R package (v1.20.0). Adjusted p-values were calculated via the Benjamini-Hochberg method, with differentially expressed genes (DEGs) defined as those exhibiting an adjusted p-value < 0.05 ^[25]^.

### 2.4 Quantitative real-time PCR

Total RNA was isolated using TRIzol reagent. Subsequently, complementary DNA (cDNA) was synthesized from the extracted RNA using the PrimeScript™ RT reagent kit (Takara Bio, Inc.). Gene expression levels of *POU5F1B* and *WNK2* were quantified by real-time quantitative PCR (qPCR) using TB Green Premix Ex Taq™ (Takara Bio, Inc.), with *GAPDH* serving as the endogenous reference gene for normalization. The primer sequences used for amplification of *WNK2*, *POU5F1B*, and *GAPDH* were as follows: WNK2, F: 5’-TGG TTC ATC ATC TGT CCG-3’and R: 5’-AAG CTG GGT TGT TCC TT-3’. POU5F1B F: 5’-GAA CCG AGT GAG AGG CAA CC-3’ and R: 5’-GAT GTG GCT GAT CTG CAG TGT-3’; GAPDH F: 5’-AGC CAC ATC GCT CAG ACAC-3’ and R: 5’-TTA AAA GCA GCC CTG GTG AC-3’.

### 2.5 Western blotting

Cell lysate buffer was prepared using radio-immunoprecipitation assay buffer, 1% protease inhibitor cocktail, and 1% phosphatase inhibitor. The protein concentration was measured using a bicinchoninic acid protein assay reagent kit. For the sample preparation, each sample was balanced to an equal volume and equal quantity. After electrophoresis, the protein was immobilized on polyvinylidene fluoride (PVDF) membranes and nonspecific protein blocking was performed, we incubated the PVDF membranes with the primary antibody at 37 ℃. The antibodies used were anti-WNK2 (cat: ab192397, 1:500, Abcam), anti-POU5F1B (cat: ab230429, 1:1,000, Abcam), anti-actin (cat#3700, 1:1,000, CST), anti-mouse (cat#7076, 1:5,000, CST), and anti-rabbit (cat#7074, 1:5,000, CST) antibody .

### 2.6 Immunohistochemistry

POU5F1B expression was evaluated in 70 OC tissues and 10 adjacent non-tumor tissues using commercial tissue microarray HOvaC070PT01 (Outdo Biotech). Immunohistochemistry (IHC) was conducted by the immunohistochemistry kit (Gene Tech Company Limited) ^[26]^. After deparaffinization at 60 ℃ for 30 mins, we retrieved the citrate antigen and blocked nonspecific antigen successively. We incubated Anti-POU5F1B antibody (cat: ab230429, 1:1,000; Abcam) at 4 overnight. After secondary antibody incubation, we stained the section with 3,3-diaminobenzidine.

### 2.7 Proliferation and transwell assays

Cell viability was measured using a cell counting kit-8 (CCK8) or colony formation assay. For the CCK8 assay, transfected cells were incubated in 96-well plates, and CCK-8 kits were used according to the manufacturer’s instructions. For colony formation, we incubated 2,000 cells in 12-well plates for 2 weeks. Subsequently, the colonies were counted by staining them with crystal violet.

Transwell chambers with 8 µm pores were used for transwell analysis. Cells (2 × 10^4^) suspended in a serum-free medium were incubated in the upper chamber. The medium containing 10% serum was placed in the lower chamber. Transwell chambers were placed in an incubator for 48 h. We then wiped off the cells in the upper chamber and fixed the migrated cells with 4% paraformaldehyde. Finally, the cells were stained with crystal violet. Rescue assays were performed to determine whether POU5F1B could rescue the effect of WNK2 on the viability and invasion of cancer cells.

### 2.8 Nude mouse xenograft model

To investigate whether WNK2 promotes OC malignancy via POU5F1B, we performed a subcutaneous xenograft tumor model. Twenty female six-week-old nude mice were obtained from Huafukang Co., Ltd. (Beijing, China). Experiments were conducted at the Animal Center of Army Medical University, with approval from its Institutional Animal Care and Use Committee (IACUC, Approval No. KY2024087). Next, stable cell lines (sh-WNK2 + Vector or sh-WNK2 + POU5F1B-OE) were suspended in PBS (1×10⁶ cells/100 µl). This suspension was injected subcutaneously into the flanks of mice. Tumor Monitoring: Once palpable tumors formed, tumor volume was measured twice weekly using calipers and calculated as Volume = (Width² × Length) / 2. Tumor growth curves were generated from these measurements. Immunohistochemistry (IHC): To confirm POU5F1B expression levels within the tumors, IHC analysis was performed on harvested tumor tissues.

### 2.8 Statistical analysis

Statistical analyses were performed using GraphPad Prism 8.0. Data are presented as mean ± SD from three independent biological replicates. Intergroup differences were assessed using Student’s t-tests (paired or unpaired, as appropriate). A *p*-value < 0.05 was considered statistically significant.

## Results

### 3.1 POU5F1B is the candidate target gene of WNK2

Our prior work established the distinct role of WNK2 in ovarian cancer (OC) compared to other malignancies. Analysis of TCGA data (GEPIA portal) confirmed lower *WNK2* expression in liver, pancreatic, glioma, gastric, and colon cancers (Figure 1a). To identify WNK2-dependent transcriptional networks, we performed RNA-seq in WNK2-knockdown OC cells. A heatmap displays the top 25 downregulated differentially expressed genes (DEGs) in si-WNK2 cells (Figure 1b). Six DEGs (*POU5F1B*, *TRIB3*, *CDC42EP1*, *HES6*, *TYMP*, and *LARGE2*) were prioritized based on established roles in promoting cancer malignancy (Figure 1c) ^[27–30]^. We validated these candidates via RT-qPCR in *WNK2*-KD CAOV3 and *WNK2*-OE A2780 cells. This confirmed POU5F1B as the most significantly regulated target of WNK2 (Figure 1d).

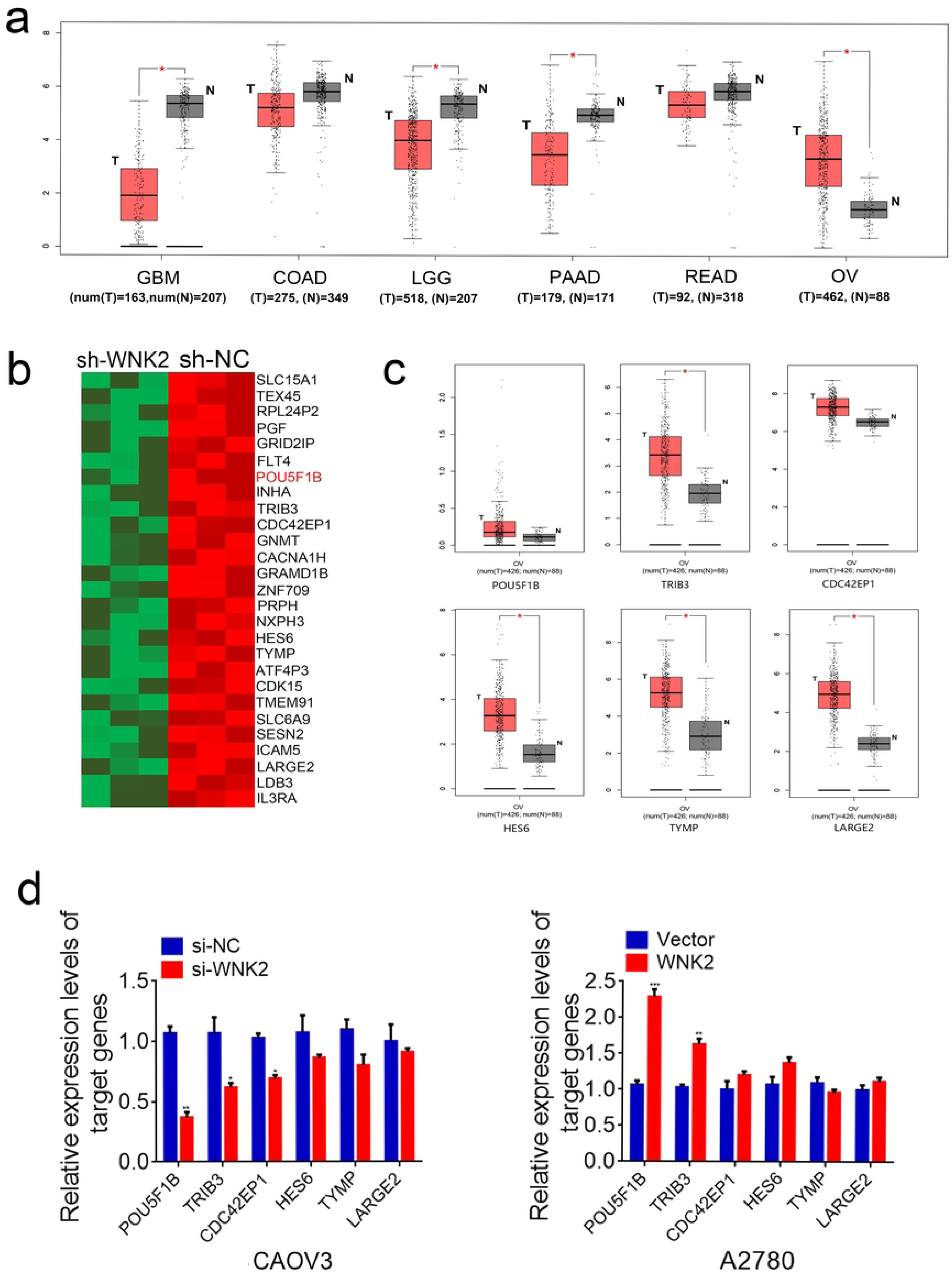

### 3.2 WNK2 regulates POU5F1B positively both in mRNA and protein levels

RT-qPCR and western blot analysis confirmed that WNK2 knockdown reduced POU5F1B mRNA and protein levels (Figure 2a, b), while WNK2 overexpression increased them (Figure 2c, d). This demonstrates that WNK2 directly and positively regulates POU5F1B expression. Based on this regulation, we investigated POU5F1B’s functional role in OC progression.

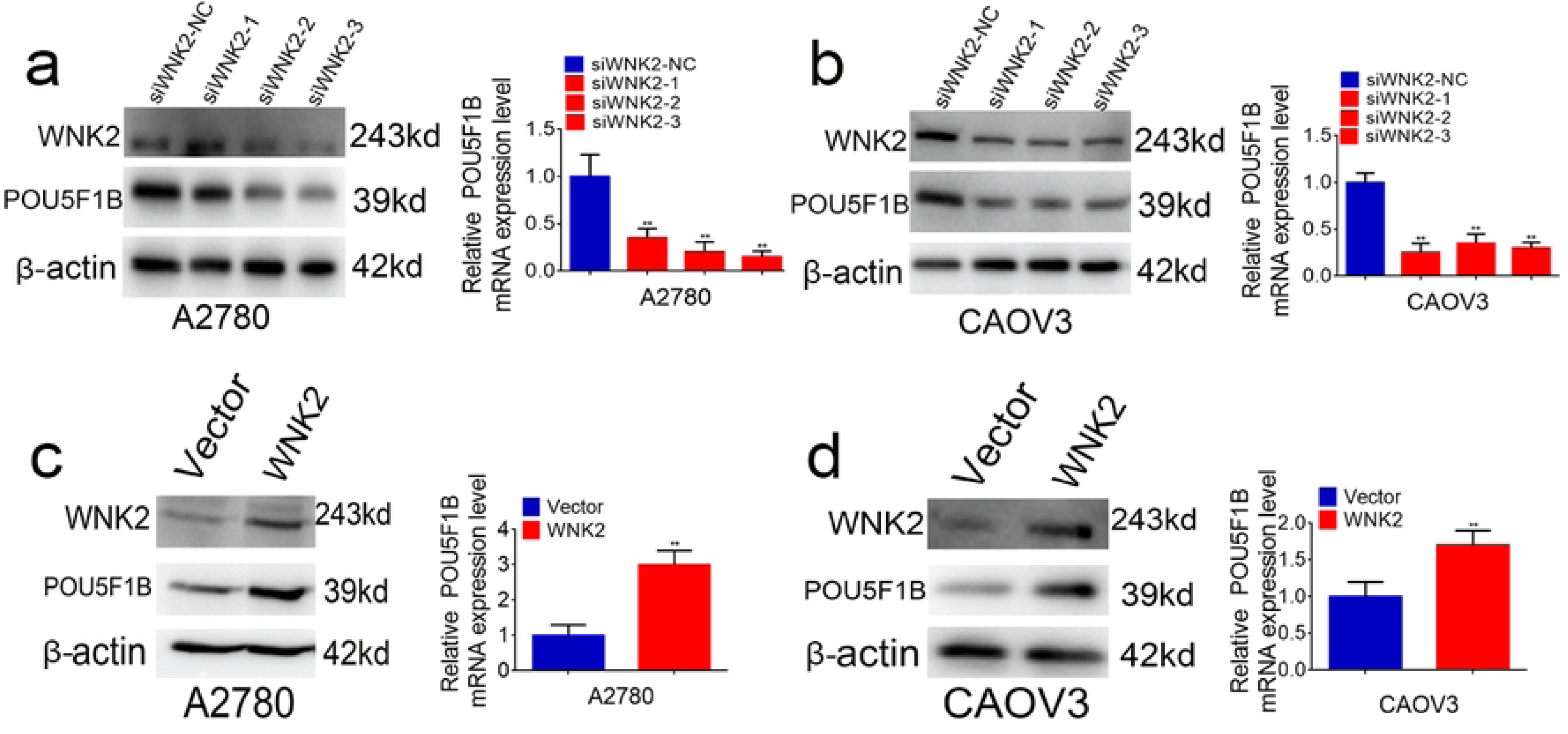

### 3.3 POU5F1B functions as an oncogene in ovarian cancer

Due to the absence of POU5F1B protein expression data in ovarian cancer (OC) within public repositories, we evaluated its expression via immunohistochemistry (IHC). Representative IHC staining demonstrates low, moderate, and high POU5F1B expression patterns (Figure 3a). Analysis of the HOvaC070PT01 tissue microarray revealed significantly elevated POU5F1B protein levels in OC tissues versus histologically confirmed adjacent normal tissues (Figure 3b, Table 1).

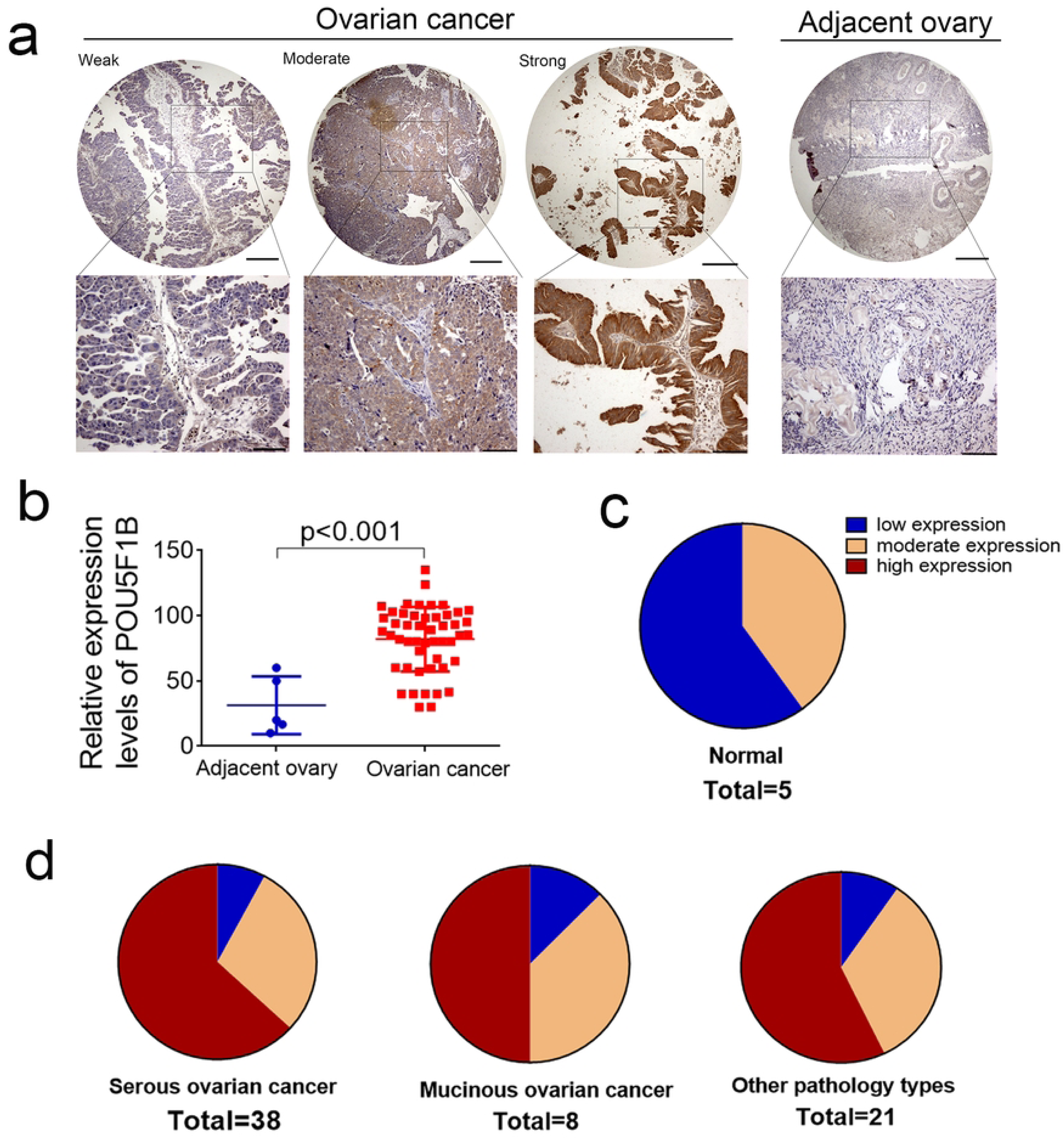

**Table 1.** POU5F1B is increased in ovarian cancer tissues compared to adjacent ovary tissues p<0.01

| Parameters | Total (n=68) |  | P-value |
| --- | --- | --- | --- |
|  | High miR-324-3p expression | Low miR-324-3p expression |  |
|  |  |  | Pathology types |
| Boderline tumors | 0 | 5 |  |
| Serous carcinoma | 24 | 14 | 0.008* |
| Mucinous carcinoma | 4 | 4 | 0.068 |
| Other pathology types | 12 | 9 | 0.024* |
|  |  |  | Ages (years) |
| ≥55 | 15 | 10 | 0.801 |
| <55 | 24 | 14 |  |

Functional experiments (e.g., CCK-8, Edu, and colony formation assays) were carried out to reveal the roles of POU5F1B in OC. When POU5F1B was successfully knocked down in A2780 and CAOV3 (Figure 4a), the proliferation of cancer cells was remarkably suppressed (Figure 4b,c). Similarly, the overexpression of POU5F1B accelerated the proliferation of OC cells (Figure 5b,c). Transwell analysis also proved that POU5F1B knockdown inhibits the migration of cancer cells (Figure 4d), and the overexpression promotes the migration of cancer cells (Figure 5d).

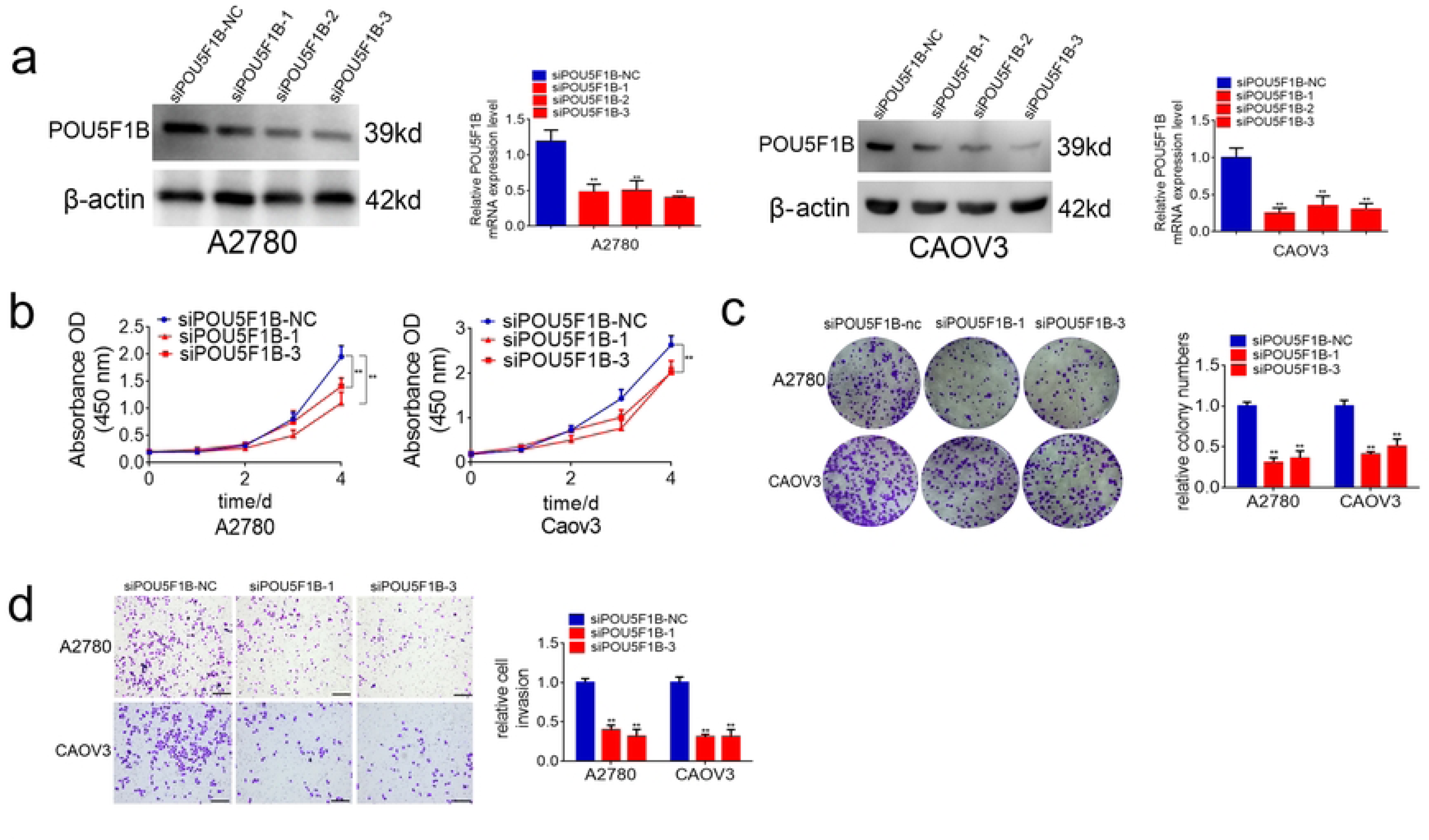

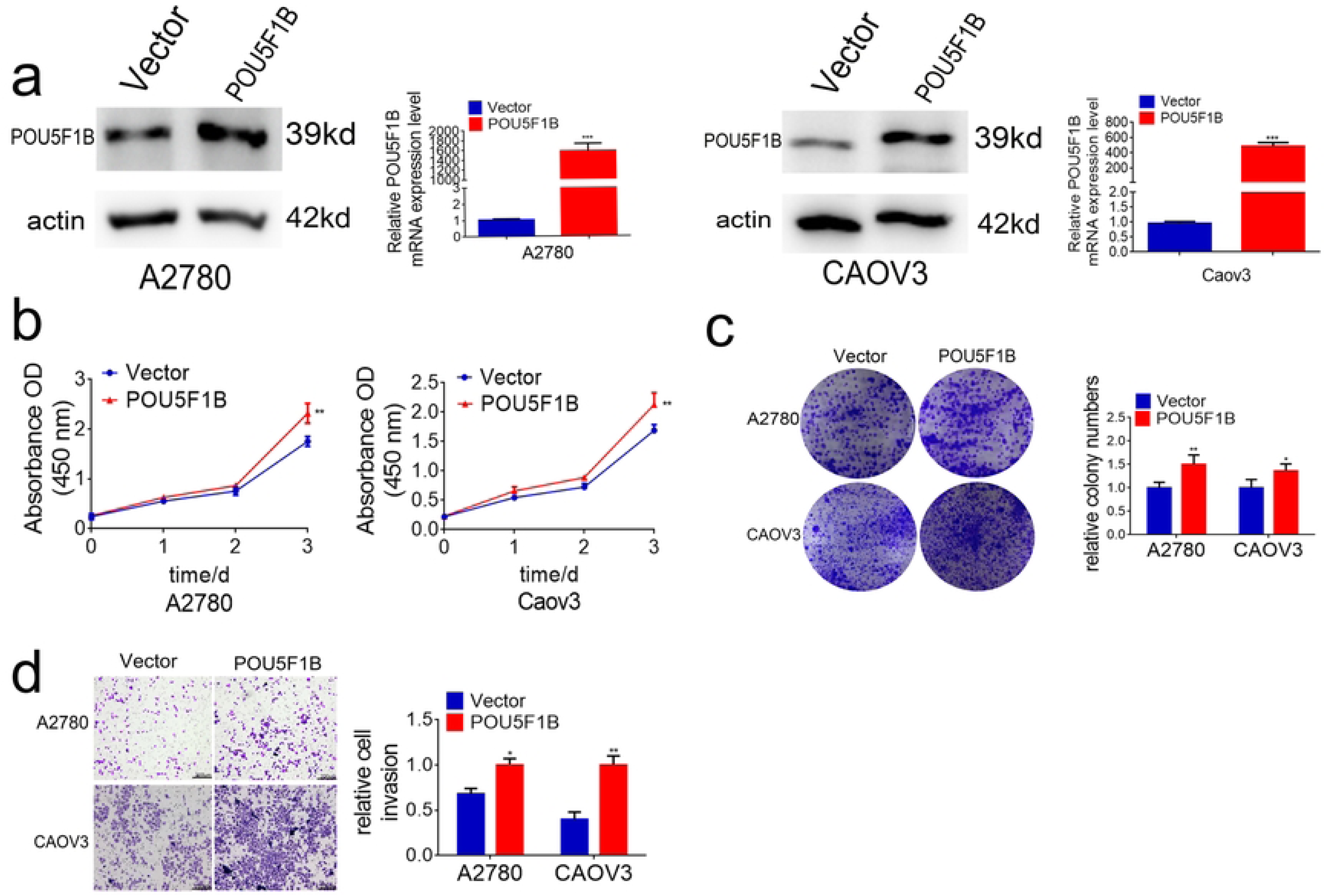

### 3.5 WNK2 promotes OC growth and activates AKT through POU5F1B

To investigate functional crosstalk between WNK2 and POU5F1B, we generated stable WNK2-knockdown (WNK2-KD) OC cell lines via lentiviral transduction, followed by POU5F1B overexpression plasmid transfection. Immunoblotting confirmed efficient WNK2 depletion and concurrent POU5F1B restoration in A2780 and CAOV3 cells (Figure 6a). POU5F1B overexpression reversed WNK2-KD-induced suppression of proliferation (CCK-8/EdU/colony formation; Fig 6b-c) and migration (Transwell; Fig 6d)^[23]^. Xenograft models demonstrated that POU5F1B restored tumor growth in WNK2-KD cells (tumor volume/weight; Fig 7a,b), with IHC confirming successful POU5F1B re-expression (Fig 7c). Given POU5F1B’s reported oncogenic function through AKT activation ^[23]^, we assessed pathway activity. WNK2-KD suppressed phospho-AKT (Ser473) without altering total AKT (Fig 8a). POU5F1B overexpression rescued phospho-AKT (Ser473) inhibition in WNK2-KD cells (Fig 8a). Based on prior phosphoproteomics identifying WNK2-mediated RAS activation ^[18]^, we inhibited RAS with salirasib:Salirasib treatment abolished WNK2-driven POU5F1B upregulation at mRNA/protein levels (RT-qPCR/WB; Fig 8b,c). This defines a WNK2-RAS-POU5F1B-AKT signaling axis in ovarian cancer.

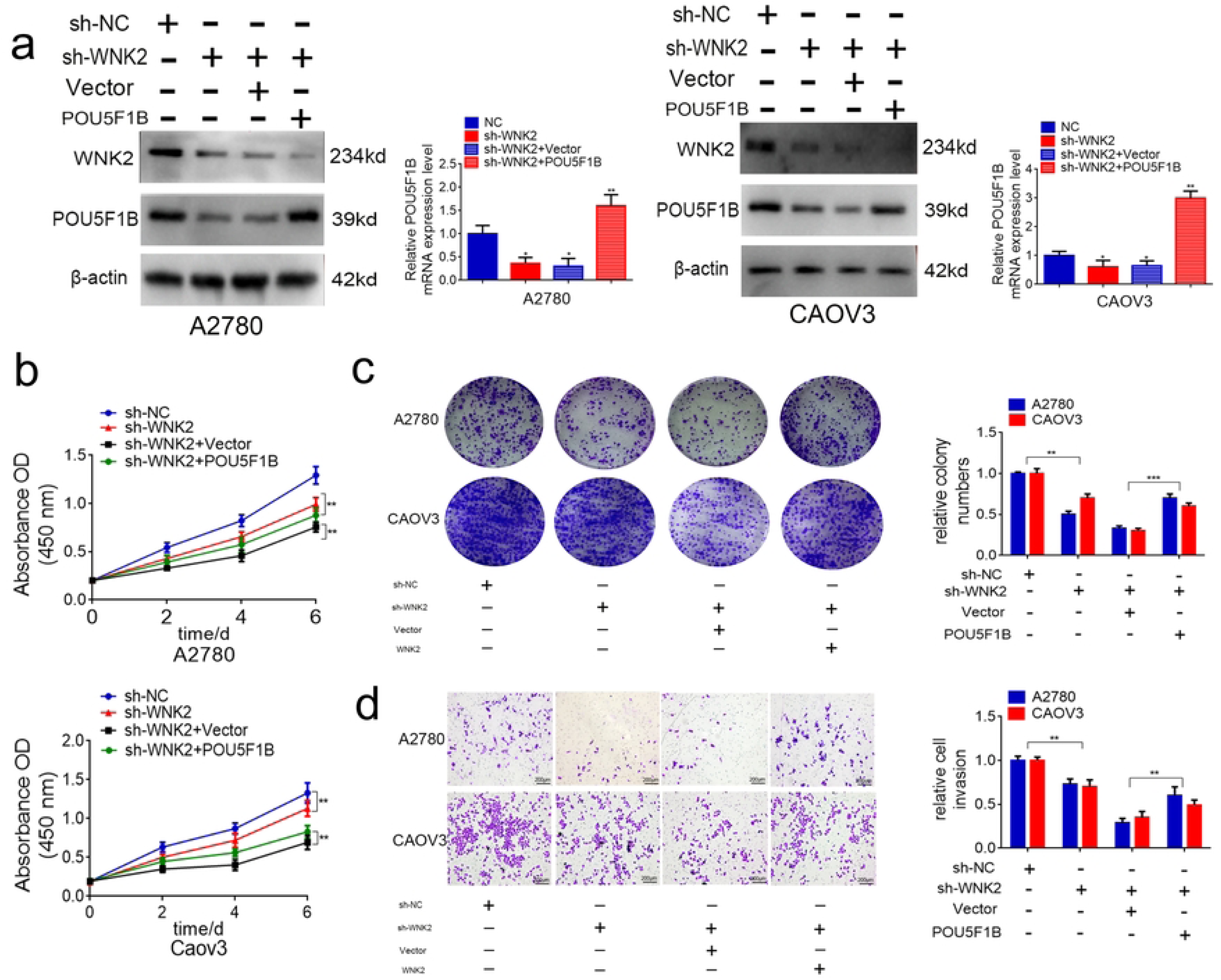

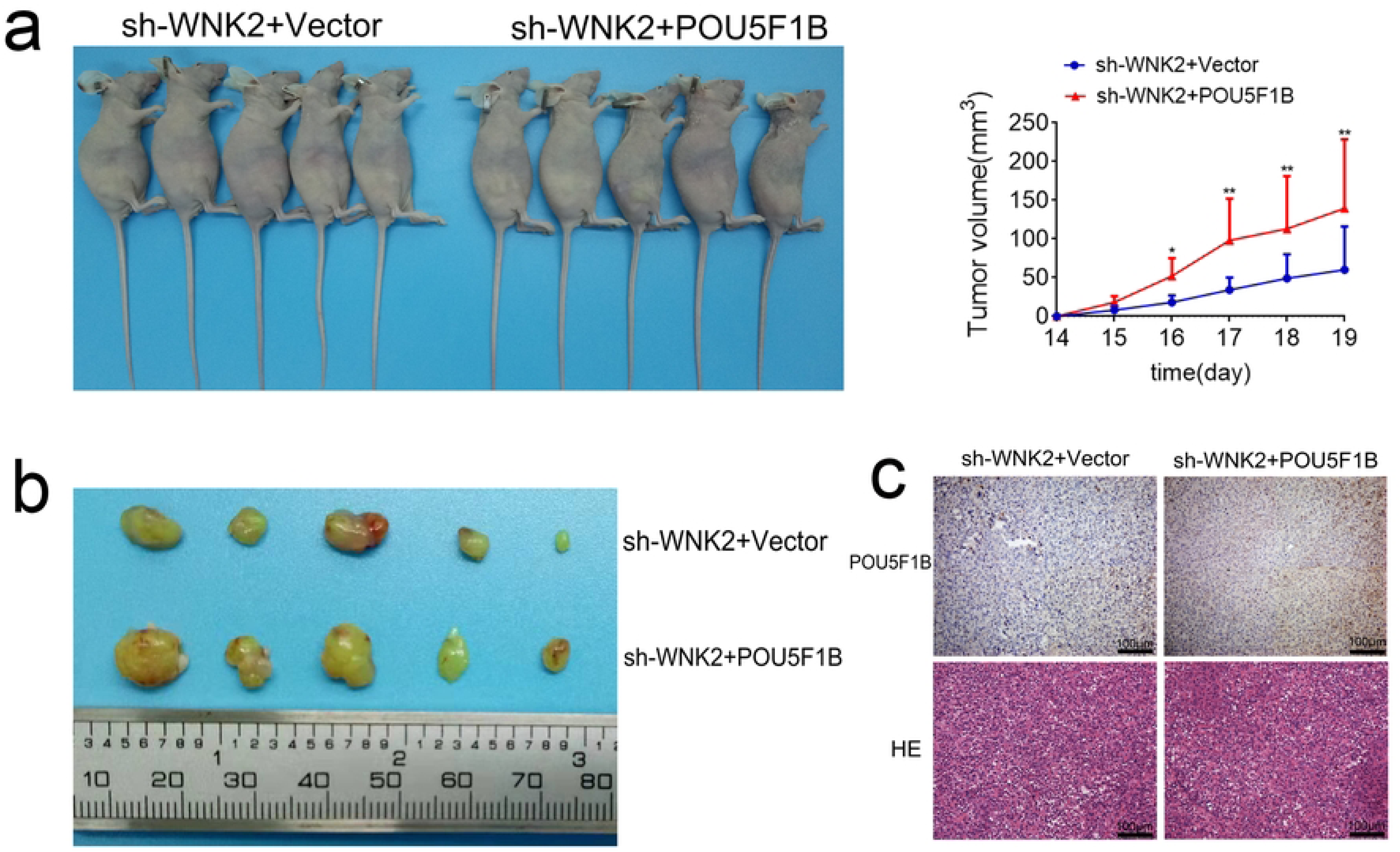

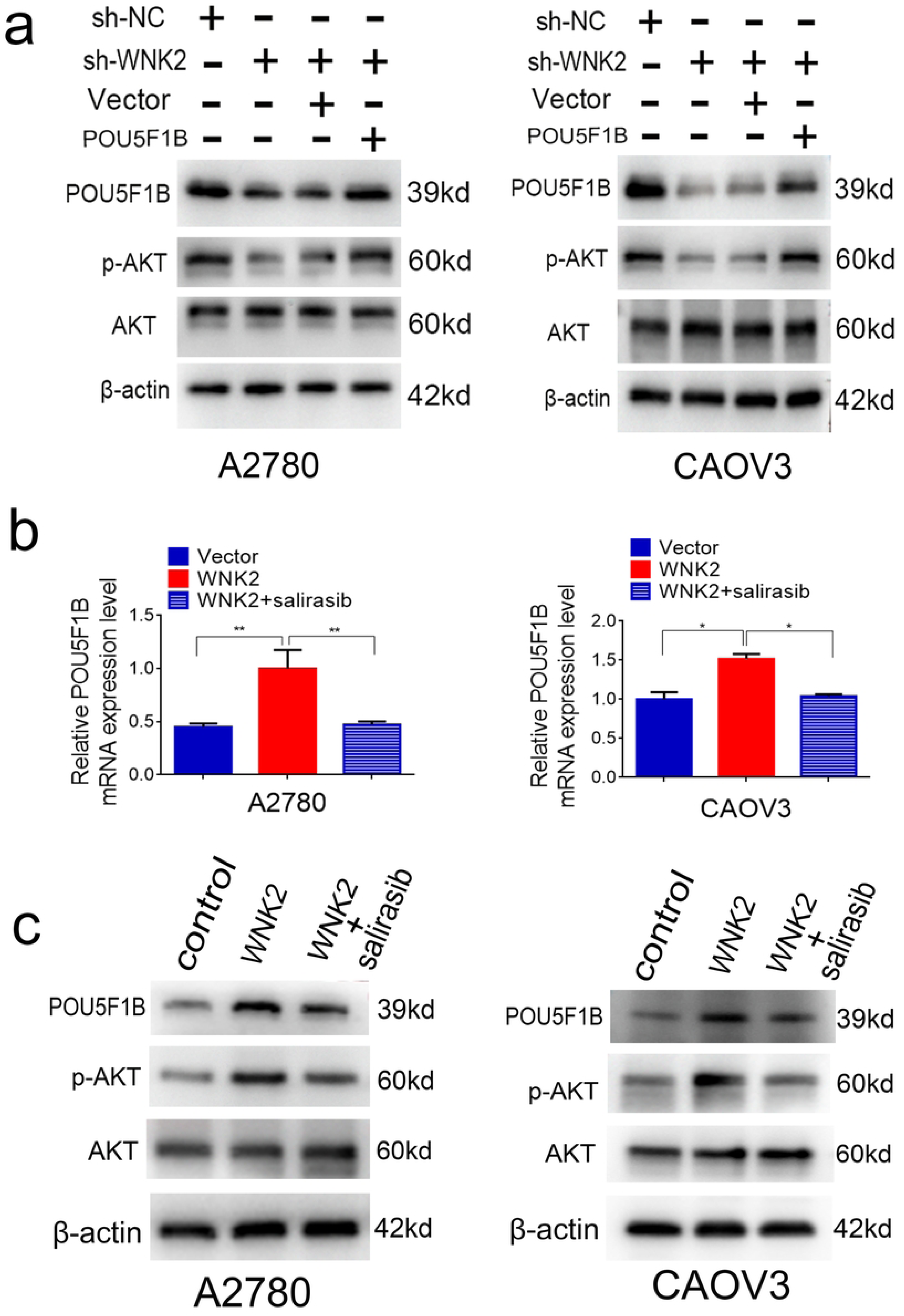

## 4. Discussion

Ovarian cancer has the highest fatality rate of all gynecological cancers, driving the urgent development of new, more effective treatments ^[31]^. The molecular mechanisms behind ovarian cancer progression, however, remain elusive. Adding a significant layer of complexity is the intricate and often counterintuitive behavior of cancer genes, exemplified by tumor suppressor genes that drive progression and oncogenes that suppress it ^[32]^. A comprehensive understanding of the contrasting functions of cancer genes is thus critical for devising effective treatments—as exemplified by WNK2, a cytoplasmic serine-threonine kinase whose structure, featuring coiled-coil domains and multiple PXXP and SH3 motifs, suggests a complex and context-dependent role ^[33]^.As a neuron-enriched kinase, WNK2 is primarily expressed in the brain and has been shown to regulate a diverse array of processes, including circadian rhythms, electrolyte homeostasis, cell survival, and proliferation^[34, 35]^.WNK2 functions as a tumor suppressor in multiple cancers, including glioblastoma, gastric, cervical, and pancreatic cancer ^[9]^. Conversely, we found that WNK2 is overexpressed in OC cells and patient tissues. This overexpression correlated with higher tumor grade and predicted poor patient survival. Functionally, WNK2 promoted OC cell proliferation and invasion in both in vitro and in vivo models. To investigate the mechanism behind this oncogenic role, we leveraged high-throughput RNA sequencing (RNA-Seq), a powerful tool that provides a comprehensive and precise transcriptomic profile ^[36]^. To identify WNK2-regulated downstream genes in OC, we performed transcriptome sequencing. Our results demonstrate that WNK2 promotes OC progression by upregulating the oncogene *POU5F1B*. As a homolog of the master pluripotency factor OCT4, *POU5F1B* possesses an intact open reading frame (ORF) with 96% amino acid identity to OCT4 isoform 1. It encodes a highly similar protein, known to interact with protein kinases and cytoskeletal molecules ^[18]^. *POU5F1B* influences cell growth and adhesion by modulating intracellular signaling pathways and the activity of trans-acting factors. Its expression is not only a prognostic marker for poor patient outcomes but also a driver of malignant transformation ^[18–21]^. In this study, we identify the oncogenic role of POU5F1B in OC for the first time, demonstrating its overexpression in OC tissues and its promotion of malignant cell behaviors. Mechanistically, we establish POU5F1B as a key downstream effector of WNK2. We show that WNK2 drives OC progression by upregulating POU5F1B, which in turn activates AKT signaling via phosphorylation at Ser473. This regulatory axis is contingent on RAS activity, as RAS inhibition abolished WNK2-mediated effects on both POU5F1B expression and AKT activation. Although these findings underscore the critical role of WNK2 in OC, its function as a kinase suggests an indirect mechanism for regulating POU5F1B mRNA, likely through an intermediary transcription factor. Identifying this factor is a primary objective for future research. Our work substantiates the targeting of the WNK2/POU5F1B/AKT axis as a promising novel therapeutic strategy for OC, a direction we are actively pursuing through drug screening efforts.

## Ethical approval

The study was conducted in accordance with the Declaration of Helsinki principles. It was approved by the Research Ethics Committee of the Shanghai Outdo Biotech Company.

## Author Contributions

Z.L. and Y.W conceived and designed this work. F.L. and Y.J. performed experiments and collected the data; F.L. wrote the paper; X.M, Y.J, P.Z., Y.L, and L.D reviewed and revised the paper.

## Data Availability

All relevant data are within the manuscript and its Supporting Information files.

NO

